# Designing a Substance Misuse Data Dashboard for Overdose Fatality Review Teams

**DOI:** 10.1101/2025.03.21.25324394

**Authors:** Marie Pisani, Madeline K. Oguss, Julia Dickson-Gomez, Constance Kostelac, Amy Parry, Starr H. Moss, Elizabeth Salisbury-Afshar, Brian W. Patterson, Michael F. Spigner, Megan Gussick, Alison Krautkramer, Tim Gruenloh, Askar Safipour Afshar, Preeti Gupta, Anoop Mayampurath, Majid Afshar

## Abstract

**Objective:** Overdose Fatality Review (OFR) is a public health process in which cases of fatal overdose are carefully reviewed to identify prevention strategies. Current OFR requires review of multiple unconnected data sources, which is a manually intensive process. We aimed to use human factors design principles to develop a comprehensive dashboard that could facilitate enhanced processes to support OFR.

**Materials and Methods:** We first surveyed OFR leaders in Wisconsin using the NASA Task Load Index (NASA-TLX) and conducted semi-structured interviews to identify targets for workflow optimization. Next, we developed a prototype dashboard for evaluation using a synthetic dataset built with Generative Pre-Trained Transformer (GPT-4). We subsequently performed iterative design sessions with heuristic evaluations, and end-user feedback on the final prototype was obtained via targeted surveys and semistructured interviews.

**Results:** The NASA-TLX revealed a moderately high mental workload with the current workflow. Interviews affirmed that technological challenges and reliance on manual processes were contributory. The prototype dashboard addressed these concerns by integrating multiple data sources to generate population-level visualizations and patient-level event timelines. End-users reported the potential for improved efficiency and data accessibility compared to antecedent processes.

**Discussion:** OFR is a data-intensive process that traditionally demands substantial manual effort. The data dashboard offers an informatics-based approach to streamline data aggregation and presentation, potentially enhancing the efficiency of case reviews.

**Conclusion:** Implementing a dashboard that consolidates and visualizes disparate data sources has the potential to alleviate the manual workload in overdose fatality review.

## BACKGROUND AND SIGNIFICANCE

Drug overdose death rates have been steadily rising over the past two decades, with the most significant annual spike occurring between 2019 and 2020, when rates surged by 31.0 % in the United States.[1] To address this public health crisis, some local health departments around the country have assembled Overdose Fatality Review (OFR) teams. OFRs are multidisciplinary and multi-agency teams with representatives from areas such as public health, safety, social services, medical examiners and coroners (ME/C), emergency responders, substance use treatment providers and other community stakeholders.[2] These teams discuss the local trends in overdose fatalities and review individual cases to identify and implement recommendations aimed at preventing overdose deaths.[2] OFR teams can help facilitate harm reduction strategies such as syringe services programs, naloxone education and training, outreach programs, and coordination of treatment services from healthcare settings.[3] However, many counties have yet to implement OFRs, and those that exist exhibit considerable heterogeneity in their data collection procedures, requiring substantial effort in data curation.

OFR teams in Wisconsin perform fatality reviews by integrating population- and individual-level data. These reviews involve an in-depth exploration of an individual’s timeline before their death, focusing on potential opportunities for intervention. The data may come from different sources, including the ME/C Office, emergency medical services (EMS), the Department of Corrections (DOC), law enforcement, social media, and other local and state agencies. Often, obtaining these data requires collecting data on individual cases from each agency and linking across multiple sources.

The Substance Misuse Data Commons (SMDC) was designed to address the issue of siloed datasets by linking them in a single cloud-based data repository, but it currently lacks an interface for data visualization.[4] Dashboards have been recognized as effective tools for visualizing public health data and facilitating disease surveillance, targeted analyses, and decision-making.[5] In several states, such as Indiana and North Carolina, health departments have used dashboards of aggregate data to monitor overdose trends and uncover preventable risk factors.[6,7] While these dashboards are effective for tracking local and state-wide trends and setting case review priorities, they often lack the breadth of data and the level of detail needed by OFR teams.

### OBJECTIVE

The objective of this study was to assess current data processes utilized by OFR teams and to design a data dashboard that leverages the SMDC to potentially streamline and support the data collection done by OFR teams. In the first phase of the study, the contemporary workflow was assessed to help establish the project’s necessity, scope, and operational requirements. During the second phase, a prototype dashboard was developed and evaluated.

## MATERIALS AND METHODS

### Phase 1: Analysis of Current Workflow

We conducted surveys and semi-structured interviews with OFR leaders in Wisconsin to document the current workflow and associated mental workload. To be included in the study, the participant was required to take part in at least one of three tasks in their current job responsibilities: (1) aggregating/analyzing population-level data; (2) selecting cases for case reviews; and/or (3) abstracting data for individual case reviews. Participants provided consent and were enrolled via a link to the survey in a secure, web-based Research Electronic Capture (REDCap) database.[8,9]

We used the validated NASA-Task Load Index (NASA-TLX) survey to assess the cognitive workload of the contemporary OFR process.[10] Participants were asked to select the OFR task(s) in which they participated. For each task, they completed the survey to evaluate workload across six dimensions: mental demand, physical demand, temporal demand, performance, effort, and frustration. The mean and standard deviation (SD) of all survey responses were calculated.

Survey participants were then invited to a 30-minute semi-structured interview with a research team member. This interview approach combined a predefined structure with flexibility for follow-up questions based on participants’ responses.[11] The interview guide focused on participants’ responsibilities, workflows, mental workload, key data sources, challenges, and reflections on group survey findings. The Wisconsin OFR training and technical assistance providers and a senior qualitative research expert (JDG) reviewed the guide to ensure its relevance and rigor. Interviews were conducted between October 2023 and January 2024 with participants providing verbal informed consent through secure, virtual conference software. The audio recording was transcribed and reviewed by the interviewer (MP).

The constant comparative method was used to analyze the transcript, and inductive coding was applied to organize the information into emergent categories.[12] The codebook was repeatedly revisited and revised during the process.[12] When no new categories emerged from the analysis of additional interviews, code saturation was determined to be met. Using the final codebook, all interviews were coded by one researcher. Three interviews were coded by a second researcher to assess intercoder reliability (ICR), exceeding the typical 10-25% double-coded interviews required to establish ICR.[13] The two researchers then compared coded segments and disagreements were adjudicated.

### Phase 2: Data Dashboard Design and Evaluation

To mitigate data privacy concerns during dashboard development, a synthetic dataset was patterned on SDMC data.[14] The synthetic data were generated using the Generative Pre-Trained Transformer (GPT-4) application programming interface (OpenAI™),[15] with chain-of-thought instructions to create each variable from the data dictionary, similar to other best practices in prompt engineering.[16] The prompt incorporated aggregate cohort demographic descriptive statistics to preserve variable distributions within the SMDC dataset. Initial prompts were tested with 20 rows of patient data to evaluate the quality of the output before a dataset of 300 patients was created. This dataset was then scaled up to 1,273 patients using YData™, a synthetic data generation software that employs Generative Adversarial Networks (GANs) to produce large volumes of data accurately replicating the statistical characteristics of the original data—in this case, the smaller synthetic dataset.[17]

An initial prototype of the dashboard was developed using Microsoft Power Business Intelligence™ (©Microsoft 2024).[18] The dashboard was designed to emulate the workflow and highlight the priority data sources identified in Phase 1. The dashboard consisted of three functional components: (1) visualizations of population-level data to identify demographics and trends **(Figure 1)**; (2) line-level data, such as individual patient timelines, to facilitate case-based reviews **(Figure 2)**; and (3) prediction tools, including census tract-level EMS patient incident predictions, de-identified hospital note topics, and a 30-day risk score for hospital readmission or death.

**Figure 1.**
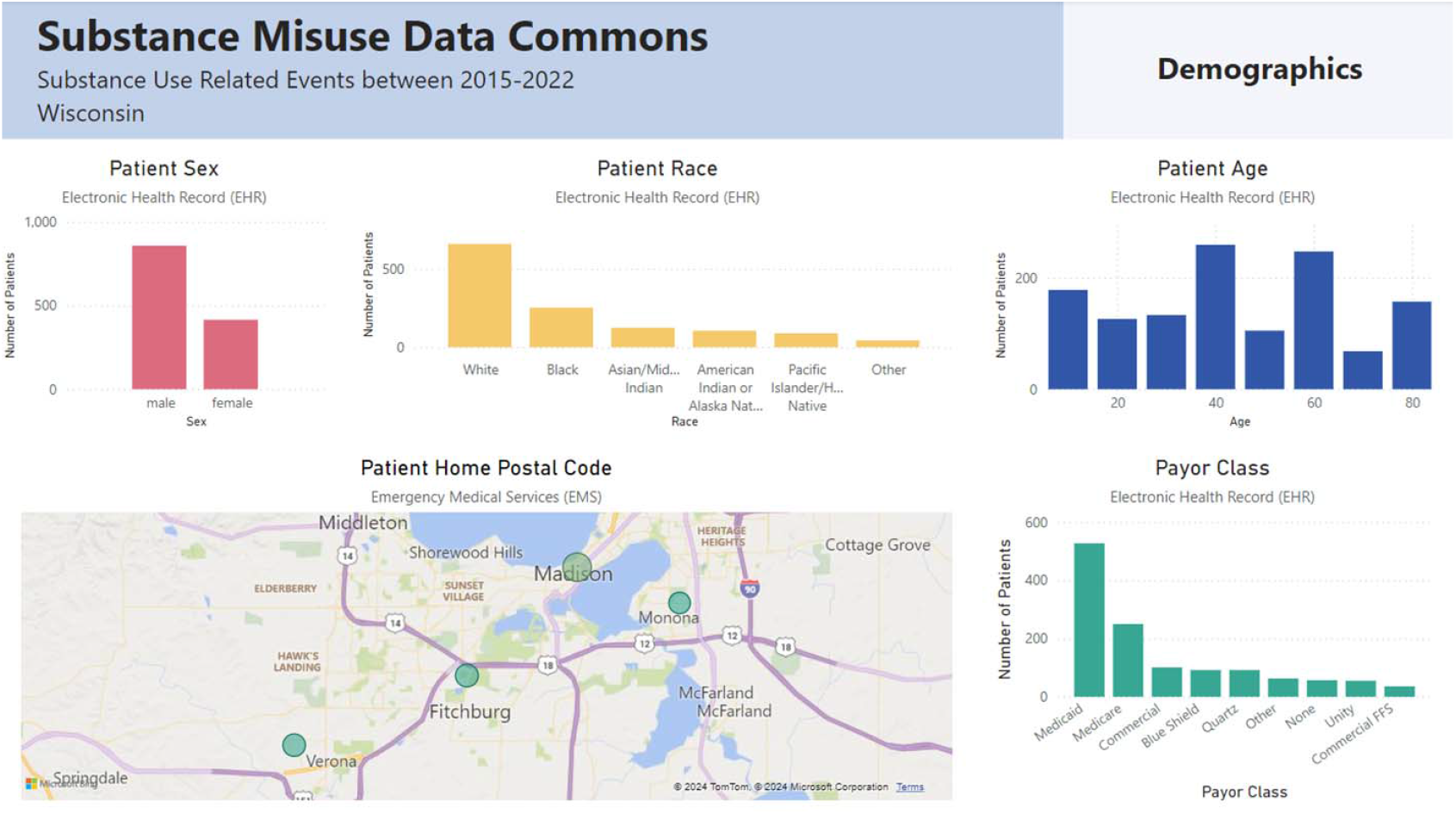
Population-level Visualizations.

**Figure 2.**
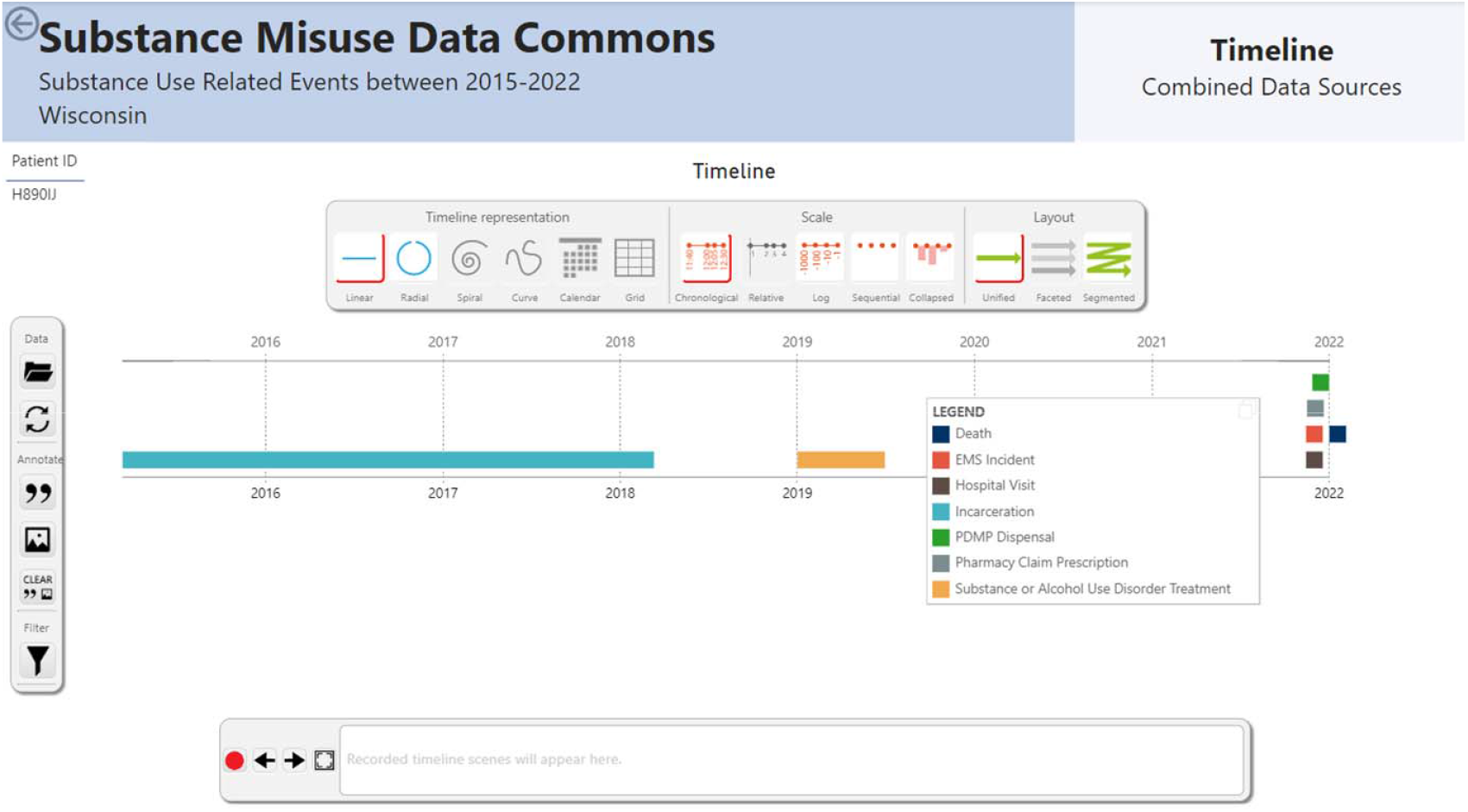
Patient Timeline Visualizations.

We conducted two iterative design sessions with emergency medicine physicians, including two EMS medical directors, a prehospital informatician, and a clinical human factors design expert. The Heuristic Evaluation Checklist for Dashboard Visualizations was used to identify and address major usability issues.[19] **(Appendix A)** These sessions refined the dashboard’s content, organization, and visual elements, culminating in a final prototype. After finalizing the prototype, a demonstration video (<u>linked here</u>) was shared with participants to showcase its content, organization, and key features.

### Phase 2: Dashboard Design and Evaluation

End-user perceptions of the final prototype were assessed with semi-structured interviews and surveys, using the same recruitment strategy and procedures as Phase 1. Two validated survey tools were used to evaluate end-user probability of adoption (Unified Theory of Acceptance and Use of Technology; UTAUT) [20] perception of usefulness (Perceived Usefulness Questionnaire) [21]. The interview guide explored participants’ perceptions of the dashboard, conditions for use, advantages, and suggested changes. One researcher coded all seven interviews using the final codebook, and a second researcher independently coded three interviews to establish inter-coder reliability (ICR). Disagreements were adjudicated. The mean and SD of all survey responses were calculated.

The full set of NASA-TLX, UTAUT and Perceived Usefulness Questionnaire questions, our interview guides and codebooks, our synthetic dataset and prompts, and our demonstration video, are all viewable in our GitLab repository.[22] The study followed the consolidated criteria for reporting qualitative research (COREQ) reporting guidelines[23] **(Appendix B)**. This research was reviewed and approved by the Institutional Review Board (IRB) at the University of Wisconsin, Madison (IRB number 2023-1091).

## RESULTS

### Phase 1: Analysis of Current Workflow

#### Surveys

Eleven OFR leaders, representing both county- and state-level agencies participated in the survey. **Table 1** provides the characteristics of the participants. The survey assessed the cognitive workload of three distinct tasks: (1) aggregating/analyzing population-level data; (2) selecting cases for case reviews; and/or (3) abstracting data for individual case reviews. Tasks had unequal sample sizes due to variations in task participation among participants. High mental workload was reported across all tasks **(Figure 3)**. The time required for each task varied among participants. Per case review period, aggregating population-level data took an average of 5.5 hours (SD 3.09), selecting cases for case review averaged 7.95 hours (SD 5.67), and abstracting data for individual case reviews required an average of 10.50 hours (SD 8.86).

**Table 1:** Phase 1 and 2 Survey Demographics.

|  | Phase 1 | Phase 2 |
| --- | --- | --- |
| Demographic Variable | Participants (n=11) | Participants (n=6) |
| Age, n (%) |  |  |
| 20-29 | 5 (45.4) | 2 (33.3) |
| 30-39 | 3 (27.3) | 0 (0.0) |
| 40-49<br>50-59 | 2 (18.2)<br>1 (9.1) | 3 (50.0)<br>1 (16.7) |
| Sex, n (%)<br>Female | 10 (90.9) | 6 (100.0) |
| Race, n (%)<br>White | 11 (100.0) | 6 (100.0) |
| Education, n (%)<br>Technical school, vocational training, community college<br>Bachelor's degree<br>Master's degree | 1 (9.1)<br>1 (9.1)<br>9 (91.8) | 0 (0.0)<br>1 (16.7)<br>5 (83.3) |
| Sector, n (%)<br>Public health<br>Government<br>Education | 9 (81.8)<br>1 (9.1)<br>1 (9.1) | 6 (100.0)<br>0 (0.0)<br>0 (0.0) |

**Figure 3.**
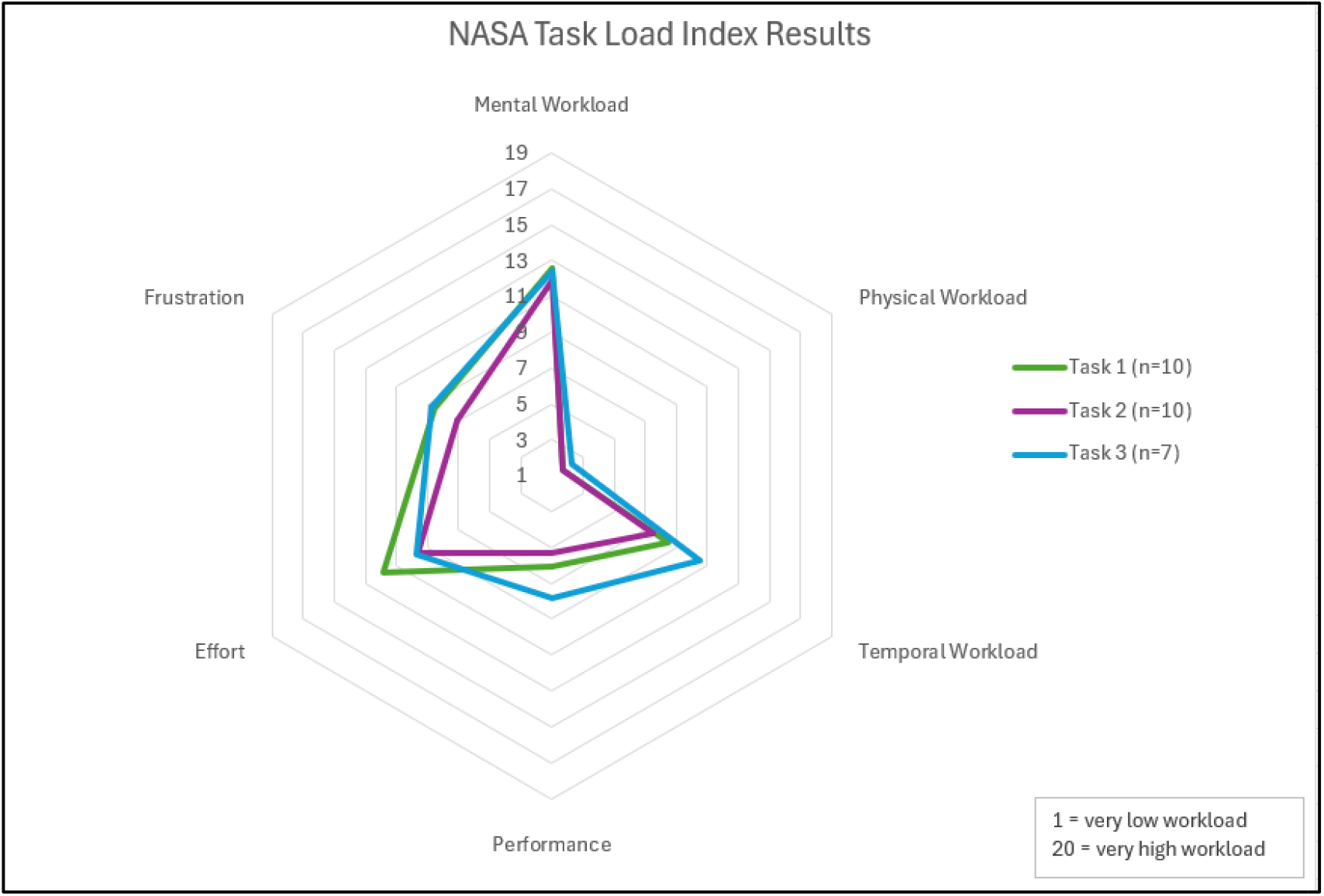
NASA-TLX Survey Results.

### Phase 1: Analysis of Current Workflow

#### Interviews

Ten of the eleven participants completed qualitative followup interviews. Participants were asked to describe their data collection process, case selection methods, challenges, desires for a future state, and key data sources. Key OFR data sources highlighted by participants are presented in **Table 2**. Summaries and selected quotes are highlighted below, while the full interview results and quotes are detailed in **Appendix C**.

**Table 2.** Key Data Sources.

| Category | Data Contributors and Resources |
| --- | --- |
| High impact data contributors | 1. State Vital Records (SVR)<br>2. Next-of-kin interviews<br>3. OFR agency partners |
| Key agency partners and contributors | 1. ME's or Coroner's Office<br>2. EMS<br>3. DOC |
|  | 4. Law enforcement |
| Other data sources | 1. Prescription Drug Monitoring Program (PDMP)<br>2. Electronic Surveillance System for the Early Notification of Community-Based Epidemics (ESSENCE)<br>3. First Watch (fire and rescue)<br>4. Wisconsin Statewide Health Information Network (WISHIN)<br>5. Overdose Detection Mapping Application Program (ODMAP)<br>6. Consolidated Court Automation Programs (CCAP)<br>7. Internet<br>8. News<br>9. Social Media |

#### Process

Participants described a typical workflow while preparing for an OFR meeting. Population-level data are analyzed to identify recent community trends. Next, representative cases are selected for review and permission is obtained from local law enforcement to review the cases. OFR leaders then coordinate the collection of available and pertinent information on the decedents across multiple sources, which may include their own sources, agency partners, and next-of-kin interviews with the decedent’s family or friends. After compiling the data, they prepare a presentation for the case review meeting, often including a timeline of the decedent’s interactions with various agencies. During the sessions, OFR members, agency partners, and community representatives review the data and collaboratively brainstorm strategies for overdose prevention.

#### Case selection

Participants reported several factors that influence case selection. Fifty percent of participants reported looking at demographic trends such as age, race, and sex. Fifty percent of participants reported combining multiple demographic and substance trends into a single theme and looked explicitly at decedents within that theme. For example, one participant stated, “Now that we’re doing theme selection, we may focus on a specific drug, like, I think the next theme that we’re doing is African American men between certain ages that historically used cocaine, but fentanyl was also involved in their cause of death.” Sixty percent prioritize selecting cases with comprehensive data, though they noted this is challenging due to limited data availability during the initial selection process. Other key factors influencing case selection included obtaining permission from agency partners (50%) and ensuring cases fell within jurisdictional boundaries (90%).

#### Case data collection and preparation

OFR leaders reported collecting data on decedents using publicly available databases and resources provided by their health departments and from the state-level data provided. They also reported requesting data from agency partners regarding any interactions with the decedent. After data are collected, they are processed manually by the OFR leader. One participant stated, “Then those individual partners have to go in, look at the specific case and then they have to like hand-put in all of the info and then they send those to me and then I scan them into our system so that we have them electronically, and then I have to take all of those electronic copies and upload those, one question at a time into REDCap.” After processing, all participants reported compiling the information into a timeline to display during case review presentations, which helped viewers to understand the decedent’s story.

#### Reported challenges

When asked about challenges, ninety percent of participants identified a reliance on manual processes to collect data as a significant challenge. Due to limited bandwidths, responses to data requests from agency representatives are often delayed or incomplete, which impacts the preparation of case review materials. When final requests were not fulfilled, critical data were missing from presentations. Participants also reported several technological challenges, including siloed data sources (40%), confusing data formatting (70%), and other technological issues (100%), all of which impacted data collection and preparation. As a result, 90% of participants highlighted time pressure as a major challenge for preparing for the OFR process

#### Desired future state

Half of the participants indicated that easier access to current data sources would be helpful, specifically mentioning simplified access as well as fewer lags to be able to identify and respond to current trends. Participants mentioned that additional data would help them. Seventy percent of participants desired healthcare data, most commonly substance use disorder treatment data; however, these data are protected by federal statute, which adds complexity to accessing and sharing them. Fifty percent of participants desired criminal justice data, whether they did not have access to it or did not often receive it when requested. Participants mentioned that they worked with multiple law enforcement agencies and regularly received data from some but not others. Additionally, two participants mentioned that they had not been able to perform next-of-kin interviews due to barriers in setting up interviews. Other desired data sources included childhood information from school districts or Child Protective Services and input from local organizations.

Eighty percent of participants mentioned that more collaboration or support would help the OFR process. Other participants specifically mentioned that increased collaboration between health departments and local agencies would help their current workflow, create the possibility to expand their services and address long-term sustainability of OFR.

### Phase 2: Data Dashboard Design and Evaluation

The synthetic data dashboard prototype, developed based on Phase 1 data, was refined into a high-fidelity prototype for further usability and human factors evaluation. The final prototype featured nine theme-based aggregate data pages covering demographics, substances used, healthcare interactions, prehospital emergency services, social and economic factors, mortality, prescription patterns, and treatment and recovery. Select components of the line-level data and timeline are shown in **Figures 1 and 2** and a full demonstration video is available in our GitLab repository.[22] The final dashboard prototype included advanced filtering capabilities, enabling users to refine data by specific time frames, substances, and death status. Three machine learning tools were integrated into the dashboard. First, the Hospital Note

Topics Tool utilized Latent Dirichlet Allocation (LDA) for topic modeling of EHR notes.[24] This tool identified prevalent themes and trends, such as patterns of substance use, healthcare utilization, and social determinants of health, providing users with a high-level understanding of key insights from unstructured text data. Second, the 30-Day Risk Score for Readmission and Death employed an XGBoost machine learning model, which analyzed a combination of EHR notes and tabular data along with EMS and neighborhood census data to predict the likelihood of hospital readmission or death within 30 days. Third, the EMS Geographic Prediction Tool combined EMS response data with neighborhood-level census tract information to identify geographic areas at higher risk for overdose events. Additionally, the final prototype incorporated a drill-down feature, allowing users to filter patient populations by category and narrow them down to individual patients.

### Phase 2: Dashboard Design and Evaluation

#### Surveys

Six OFR organizers participated in the Phase 2 survey, representing stakeholders from county, state, and federal agencies. Additional demographic details are provided in **Table 1**. All six participants reported analyzing aggregate data and selecting review cases, while four were involved in abstracting data for case reviews. UTAUT results indicated that the dashboard would likely be adopted if made available to participants **(Figure 4)**, with a mean of 4.07 out of 5.00. The Perceived Usefulness Questionnaire results suggested a moderately positive perception of usefulness for the aggregate and individual-level data **(Figure 5)**, with means of 3.61 and 3.64 out of 5.00 respectively.

**Figure 4.**
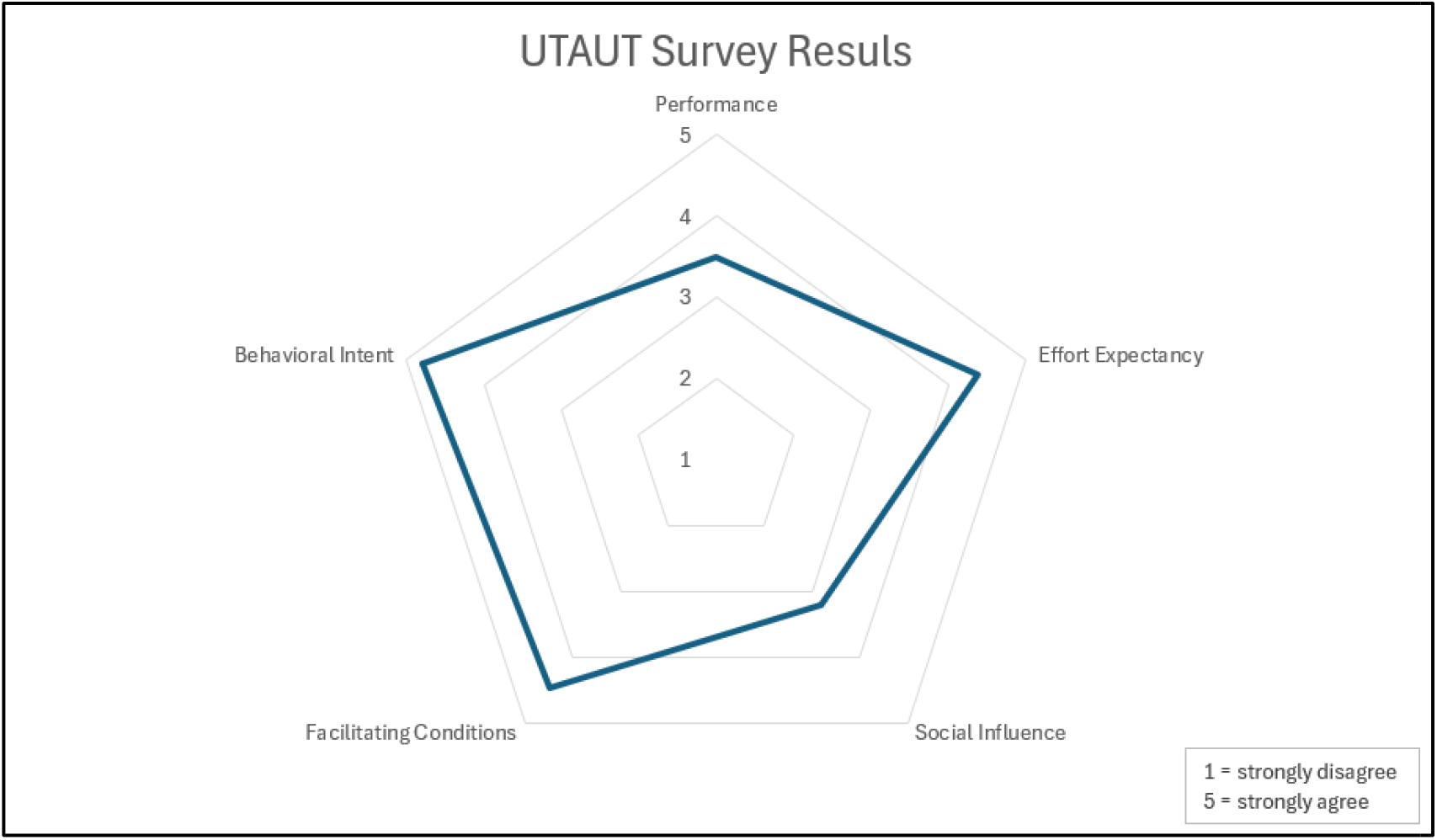
UTAUT Survey Results.

**Figure 5.**
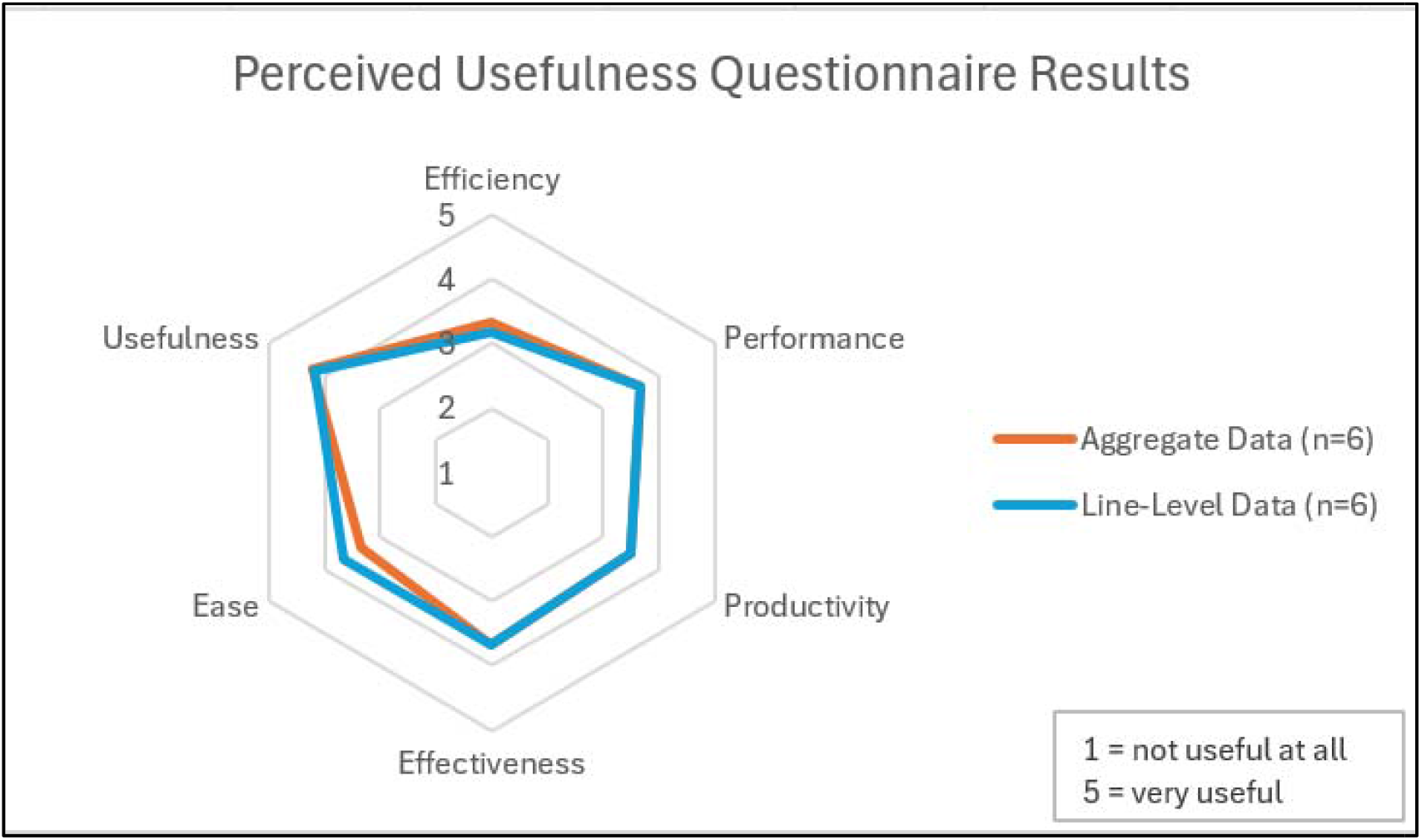
Perceived Usefulness Questionnaire Results.

### Phase 2: Dashboard Design and Evaluation

#### Interviews

Seven OFR leaders participated in semi-structured interviews. Participants were specifically asked about potential benefits of the dashboard as well as areas for improvement. Summaries of the findings are outlined below, with detailed results and quotes provided in **Appendix D**.

#### Benefits of the Dashboard

All participants highlighted improved data access as a key benefit of the dashboard. This included expanding access to currently unavailable data sources, facilitating quicker and easier access to existing data, and increasing access for less-resourced communities. Additionally, 86% of participants indicated that the dashboard could help to optimize workflows by reducing the manual processes and introducing time-saving features. Seventy-one percent of participants noted that the dashboard’s tools are especially valuable for organizational outreach efforts. The machine learning tools, particularly the EMS geographic prediction tool and the 30-day risk score for readmission or death, were highlighted as the most useful for these initiatives. All participants reported that the dashboard appeared easy to navigate, with an intuitive structure and organization. Seventy-one percent of participants specifically mentioned the ability to filter patients for specific populations as a standout feature, distinguishing the dashboard from other available tools.

#### Areas for Improvement

Most participants suggested adding more data sources. Specific recommendations included sexual orientation and gender identity, medical comorbidities such as chronic pain, and presenting data as rates instead of counts to better represent minority group trends. Concern about the accuracy and timeliness of data was expressed by 71% of participants. The SMDC cohort, which is limited to patients with hospital encounters linked to other data, excludes individuals seen only by EMS or those without EMS or hospital contact. Current data was also identified as a critical need for the dashboard. Participants proposed several technological enhancements to improve the dashboard’s usability. Specifically, 43% suggested the ability to export data from the dashboard, while 57% recommended adding other technological features such as additional filters and hover-over tips for ease of use. Other important but less commonly cited concerns included a training or learning curve (43%), having jurisdictional access (29%), adding to stigma or bias (29%), having too many years aggregated in the dataset to be reflective of current trends (29%), and the dashboard data being de-identified and therefore not being able to be connected to data about individuals from other sources (29%).

## DISCUSSION

Our study is the first to demonstrate the high cognitive workload associated with aggregation and organization of data for OFR activities. To address these challenges, we leveraged the data from the SMDC to integrate and visualize information from multiple disparate data sources, an approach not previously applied in this context. We modeled our dashboard based on real-world workflows, incorporating predictive modeling and machine learning tools to augment decision-making. Along with theme-based aggregate data pages, our dashboard includes patient-level timelines, offering a novel comprehensive and actionable view. Evaluative feedback highlighted the potential of this approach to streamline preparation for OFR meetings and identified areas for further refinement.

Our SMDC data dashboard serves as a comprehensive tool for extracting, transforming, and visualizing overdose data. It was designed to refresh with recent case information from multiple sources, incorporates automation for case matching, data formatting, and quality checks, and offers easy navigation to streamline current workflows. The dashboard helps automate the current workflow challenge of siloed data sources and includes additional data that participants found valuable. It integrates key data variables from multiple agencies, with options to filter the data by important demographic and substance-related factors. The automated timeline feature compiles data from all sources in our dataset, visually representing the events leading to an overdose death. Phase 2 participants saw the dashboard’s potential to reduce time pressure and reliance on manual processes. Additionally, in larger cities where reviewing every case is impractical or impossible, OFR leaders typically hand-pick cases to analyze in detail. By linking all cases and presenting data in aggregate, this system enhances the understanding of overall trends and helps to better inform recommendations.

Many participants indicated that they would use this dashboard as an additional tool, rather than replacing their current methods. This likely explains our Phase 2 survey findings, which revealed a discrepancy between the tool’s effectiveness and efficiency. This preference highlights a key concern raised in Phase 2 interviews that cannot be resolved in future dashboard versions: the deidentified nature of the SMDC data prevents integration with other data sources, such as next-of-kin interviews, a crucial part of the case review process. Other areas of improvement discussed in Phase 2 interviews can be resolved. Future work will refine the dashboard by adding suggested variables and technological features. We also plan to evaluate data accuracy and representativeness. Because our dashboard is currently populated with synthetic data, we plan to analyze the actual hospital-centered SMDC data and compare it to the EMS-centered OFR data.

Phase 1 participants emphasized a desire for increased collaboration and support. Some mentioned this in the form of improved relationships between the health department and other agencies. Expanding the sectors involved in OFR processes and broadening the information available for case reviews may highlight previously unseen gaps in care. ED utilization is common among those who misuse opioids and other drugs, and the number of ED visits is associated with an increased risk of drug overdose.[25] Our dashboard aims to establish and improve data sharing between OFR teams and healthcare systems, which has been identified as an important prevention strategy implementation.[26]

OFRs take place in many counties throughout the state. There is a standardized process for all teams to receive support and training and request data at the state level from some sources such as EMS and the PDMP. However, access to other data sources such as the ME/C office and public safety agencies is less consistent. To access these data, each county is required to independently set up its own data collection processes and build relationships with local and state agencies. This highlights the need to keep working toward more standardized and consistent OFR processes across the state, which the data dashboard could provide.

### Limitations

The general process reported by participants in the interviews is mainly consistent with the process in the OFR Practitioner’s Guide and the Public Health and Safety Team (PHAST) toolkit, which are guiding frameworks for health departments when creating OFRs and holding case review meetings.[27] However, the feedback in this study was collected from local OFR leaders in Wisconsin, and therefore, these findings may not be generalizable to other systems. Limitations inherent to the SMDC include its inclusion criteria and its de-identified nature. Finally, addressing challenges in the OFR process with new technology may not ultimately lead to better outcomes. The opioid epidemic is an evolving problem that will require constant evaluation and innovation to confront.

## CONCLUSION

The existing OFR process is built on a thorough, team-based approach, but it includes several cognitively demanding tasks, and there are multiple challenges to timely data preparation. Increased collaboration, access to standard, centralized tools, and comprehensive data could build upon the rigorous work already being done by OFR teams in order to further augment and automate workflows to reduce manual work. We designed a user-centered data dashboard to help reduce the cognitive workloads identified from surveys and incorporate the desired data sources and workflows gathered from the interviews. Evaluative feedback indicated many potential benefits as well as some areas for improvement. This insight will guide the development of a real-time data dashboard accessible to OFR leaders in their review process.

## Supporting information

ONLINE DATA SUPPLEMENT - Appendix A: The Heuristic Evaluation Checklist for Dashboard Visualizations

ONLINE DATA SUPPLEMENT - Appendix B: COREQ Reporting Guidelines

ONLINE DATA SUPPLEMENT - Appendix C: Phase 1 Interview Results

ONLINE DATA SUPPLEMENT - Appendix D: Phase 2 Interview Results

## Data Availability

All data produced in the present study are available upon reasonable request to the authors. The full set of NASA-TLX, UTAUT and Perceived Usefulness Questionnaire questions, our interview guides and codebooks, our synthetic dataset and prompts, and our demonstration video, are all available online at https://git.doit.wisc.edu/smph-public/dom/uw-icu-data-science-lab-public/substance-misuse-data-commons-dashboard

## ACKNOWLEDGEMENTS

This work was supported by the National Institutes of Health, National Institute on Drug Abuse [R01DA051464] and the Department of Medicine, of the University of Wisconsin School of Medicine and Public Health [233-AAM7146]. The content is solely the responsibility of the authors and does not necessarily represent the official views of the UW-Madison Department of Medicine.

## APPENDIX

**Appendix A: The Heuristic Evaluation Checklist for Dashboard Visualizations**

**Appendix B: COREQ Reporting Guidelines**

**Appendix C: Phase 1 Interview Results**

**Appendix D: Phase 2 Interview Results**

