## Supplementary material for "Designing a Substance Misuse Data Dashboard for Overdose Fatality Review Teams": ONLINE DATA SUPPLEMENT - Appendix A: The Heuristic Evaluation Checklist for Dashboard Visualizations

### Heuristic Evaluation Checklist for Dashboard Visualizations

### 1. Visibility of System Status

The system should always keep user informed about what is going on through appropriate feedback within reasonable time.

**I. Please check your response for the individual items related to this usability factor:**

| # | Usability Factor | Response | Comments |
|---|---|---|---|
| 1.1 | Does every screen have a title or header that describes its contents? | ☐ Yes ☐ No ☐ N/A | |
| 1.2 | Is there a consistent icon design scheme and stylistic treatment across the system? | ☐ Yes ☐ No ☐ N/A | |
| 1.3 | Is there visual feedback in menus or dialog boxes about which choices are selectable? | ☐ Yes ☐ No ☐ N/A | |
| 1.4 | Is there a clear indication of the current location? | ☐ Yes ☐ No ☐ N/A | |
| 1.5 | Is the menu-naming terminology consistent with the users' task domain? | ☐ Yes ☐ No ☐ N/A | |
| 1.6 | Does the system provide visibility: that is, by looking, can the user tell the state of the system and the alternatives for action? | ☐ Yes ☐ No ☐ N/A | |

**II. Please circle the overall severity rating for this usability factor:**

| No Usability Problem | Cosmetic Problem Only | Minor Usability Problem | Major Usability Problem | Usability Catastrophe |
|---|---|---|---|---|
| 0 | 1 | 2 | 3 | 4 |

**III. If you have other comments, please specify:**

### 2. Match between System and the Real World

The system should speak the user's language, with words, phrases, and concepts familiar to the user, rather than system-oriented terms. Follow real-world conventions, making information appear in a natural and logical order.

**I. Please check your response for the individual items related to this usability factor:**

| # | Usability Factor | Response | Comments |
|---|---|---|---|
| 2.1 | Are icons concrete and familiar? | ☐ Yes ☐ No ☐ N/A | |
| 2.2 | Are the section headings and subsections in each screen ordered in the most logical way? | ☐ Yes ☐ No ☐ N/A | |
| 2.3 | Is there a natural sequence to the menu choices for a data item? | ☐ Yes ☐ No ☐ N/A | |
| 2.4 | Do the selected colors correspond to common expectations about color codes? | ☐ Yes ☐ No ☐ N/A | |
| 2.5 | Are the words/concepts and phrases used in each screen familiar to users? | ☐ Yes ☐ No ☐ N/A | |

**II. Please circle the overall severity rating for this usability factor:**

| No Usability Problem | Cosmetic Problem Only | Minor Usability Problem | Major Usability Problem | Usability Catastrophe |
|---|---|---|---|---|
| 0 | 1 | 2 | 3 | 4 |

**III. If you have other comments, please specify:**

### 3. User Control and Freedom

Users should be free to select and sequence tasks (when appropriate), rather than having the system do this for them. Users will need a clearly marked "emergency exit" to leave the unwanted state without having to go through an extended dialogue. Users should make their own decisions regarding the costs of exiting current work.

**I. Please check your response for the individual items related to this usability factor:**

| # | Usability Factor | Response | Comments |
|---|---|---|---|
| 3.1 | Is there a clear exit on each document screen? | ☐ Yes ☐ No ☐ N/A | |
| 3.2 | Are all screens accessible across the system? | ☐ Yes ☐ No ☐ N/A | |
| 3.3 | Is there an "undo" function? | ☐ Yes ☐ No ☐ N/A | |
| 3.4 | Do users have the option of either clicking on menu items with a mouse or using a touchscreen/stylus? | ☐ Yes ☐ No ☐ N/A | |
| 3.5 | Can users easily move forward and backward between screens? | ☐ Yes ☐ No ☐ N/A | |

**II. Please circle the overall severity rating for this usability factor:**

| No Usability Problem | Cosmetic Problem Only | Minor Usability Problem | Major Usability Problem | Usability Catastrophe |
|---|---|---|---|---|
| 0 | 1 | 2 | 3 | 4 |

**III. If you have other comments, please specify:**

**4. Consistency and Standards**

Users should not have to wonder whether different words, situations, or actions mean the same thing.

**I. Please check your response for the individual items related to this usability factor:**

| # | Usability Factor | Response | Comments |
|---|---|---|---|
| 4.1 | Have formatting standards been followed consistently in all screens within the system? | ☐ Yes ☐ No ☐ N/A | |
| 4.2 | Are there salient visual cues to identify the active screen? | ☐ Yes ☐ No ☐ N/A | |
| 4.3 | Are there no more than four to seven colors and are they far apart along the visible spectrum? | ☐ Yes ☐ No ☐ N/A | |
| 4.5 | Are names consistent, both within each tab and across the system, in position, in grammatical style and terminology? | ☐ Yes ☐ No ☐ N/A | |
| 4.6 | Are similar procedures used to access options? | ☐ Yes ☐ No ☐ N/A | |
| 4.7 | Is color coding consistent throughout the system? | ☐ Yes ☐ No ☐ N/A | |

**II. Please circle the overall severity rating for this usability factor:**

| No Usability Problem | Cosmetic Problem Only | Minor Usability Problem | Major Usability Problem | Usability Catastrophe |
|---|---|---|---|---|
| 0 | 1 | 2 | 3 | 4 |

**III. If you have other comments, please specify:**

**5. Recognition rather than Recall**

Make objects, actions, and options visible. The user should not have to remember information from one part of the dialogue to another. Instructions for the use of the system should be visible or easily retrievable whenever appropriate.

**I. Please check your response for the individual items related to this usability factor:**

| # | Usability Factor | Response | Comments |
|---|---|---|---|
| 5.1 | Are prompts, cues, and messages placed where the eye is likely to be looking on the screen? | ☐ Yes ☐ No ☐ N/A | |
| 5.2 | Is white space used to create symmetry and lead the eye in the appropriate direction? | ☐ Yes ☐ No ☐ N/A | |
| 5.3 | Have items been grouped into logical zones, and have headings been used to distinguish between zones? | ☐ Yes ☐ No ☐ N/A | |
| 5.4 | Is color highlighting used to get the user's attention? | ☐ Yes ☐ No ☐ N/A | |

**II. Please circle the overall severity rating for this usability factor:**

| No Usability Problem | Cosmetic Problem Only | Minor Usability Problem | Major Usability Problem | Usability Catastrophe |
|---|---|---|---|---|
| 0 | 1 | 2 | 3 | 4 |

**III. If you have other comments, please specify:**

**6. Flexibility and Efficiency of Use**

The system should offer users a number of options when it comes to finding content. Users should be able to achieve their goals in an efficient manner. Also reflects the means available to the users to customize the interface, to take account of their working strategies and/or habits. It reflects the capacity for the interface to adapt to users' needs.

**I. Please check your response for the individual items related to this usability factor:**

| # | Usability Factor | Response | Comments |
|---|---|---|---|
| 6.1 | Is navigation between screens simple and visible? | ☐ Yes ☐ No ☐ N/A | |
| 6.2 | If the system uses a pointing device, do users have the option of either clicking on fields or using a touchscreen/stylus? | ☐ Yes ☐ No ☐ N/A | |
| 6.3 | On menus, do users have the option of either clicking directly on a menu item or using a touchscreen/ stylus? | ☐ Yes ☐ No ☐ N/A | |
| 6.4 | Do the users have the ability to control display configurations? | ☐ Yes ☐ No ☐ N/A | |
| 6.5 | Can the users enter default or baseline ranges? | ☐ Yes ☐ No ☐ N/A | |
| 6.6 | Can the user remove or hide unnecessary displays? | ☐ Yes ☐ No ☐ N/A | |
| 6.7 | Can the user filter information to adjust rapidly to the focus of interest? | ☐ Yes ☐ No ☐ N/A | |

**II. Please circle the overall severity rating for this usability factor:**

| No Usability Problem | Cosmetic Problem Only | Minor Usability Problem | Major Usability Problem | Usability Catastrophe |
|---|---|---|---|---|
| 0 | 1 | 2 | 3 | 4 |

**III. If you have other comments, please specify:**

**7. Aesthetic and Minimalist Design/Remove the Extraneous (Ink)**

Dialogues should not contain information which is irrelevant or rarely needed. Every extra unit of information in dialogue competes with the relevant units of information and diminishes their relative visibility. Present the largest amount of data with the least amount of ink.

**I. Please check your response for the individual items related to this usability factor:**

| # | Usability Factor | Response | Comments |
|---|---|---|---|
| 7.1 | Is only (and all) information essential to decision making displayed on the screen? | ☐ Yes ☐ No ☐ N/A | |
| 7.2 | Have large objects, bold fonts, and simple areas been used to distinguish sections? | ☐ Yes ☐ No ☐ N/A | |
| 7.3 | Are field labels brief, familiar, and descriptive? | ☐ Yes ☐ No ☐ N/A | |
| 7.4 | Is the visual layout well designed? | ☐ Yes ☐ No ☐ N/A | |
| 7.5 | Are there any unnecessary data elements in each screen? | ☐ Yes ☐ No ☐ N/A | |
| 7.6 | Is the data presented in a simple format? | ☐ Yes ☐ No ☐ N/A | |
| 7.7 | Is there white space between color representations? | ☐ Yes ☐ No ☐ N/A | |

**II. Please circle the overall severity rating for this usability factor:**

| No Usability Problem | Cosmetic Problem Only | Minor Usability Problem | Major Usability Problem | Usability Catastrophe |
|---|---|---|---|---|
| 0 | 1 | 2 | 3 | 4 |

**III. If you have other comments, please specify:**

**8. Spatial Organization**

Relates to the overall layout of a visual representation and refers to how easy it is to locate an information element in the display and the distribution of elements in representations.

**I. Please check your response for the individual items related to this usability factor:**

| # | Usability Factor | Response | Comments |
|---|---|---|---|
| 8.1 | Are all information elements clear and visible? | ☐ Yes ☐ No ☐ N/A | |
| 8.2 | Does the information follow a "logical" organization? | ☐ Yes ☐ No ☐ N/A | |
| 8.3 | Does the information provide detail on the context and detail associated with the data element? | ☐ Yes ☐ No ☐ N/A | |

**II. Please circle the overall severity rating for this usability factor:**

| No Usability Problem | Cosmetic Problem Only | Minor Usability Problem | Major Usability Problem | Usability Catastrophe |
|---|---|---|---|---|
| 0 | 1 | 2 | 3 | 4 |

**III. If you have other comments, please specify:**

**9. Information Coding**

Relates to the symbols or representations used to aid perception.

**I. Please check your response for the individual items related to this usability factor:**

| # | Usability Factor | Response | Comments |
|---|---|---|---|
| 9.1 | Are symbols appropriate for the data represented? | ☐ Yes<br>☐ No<br>☐ N/A | |
| 9.2 | Are realistic characteristics used to represent data or information elements? | ☐ Yes<br>☐ No<br>☐ N/A | |

**II. Please circle the overall severity rating for this usability factor:**

| No Usability Problem | Cosmetic Problem Only | Minor Usability Problem | Major Usability Problem | Usability Catastrophe |
|---|---|---|---|---|
| 0 | 1 | 2 | 3 | 4 |

**III. If you have other comments, please specify:**

**10. Orientation**

Provision of support for the user to orientate them in the visualization.

**I. Please check your response for the individual items related to this usability factor:**

| # | Usability Factor | Response | Comments |
|---|---|---|---|
| 10.1 | Are measurement units displayed clearly? | ☐ Yes<br>☐ No<br>☐ N/A | |
| 10.2 | Are there labels associated with each data field? | ☐ Yes<br>☐ No<br>☐ N/A | |
| 10.3 | Can the user control the level of detail they see in a representation? | ☐ Yes<br>☐ No<br>☐ N/A | |
| 10.4 | Can the user redo/ undo their actions? | ☐ Yes<br>☐ No<br>☐ N/A | |

**II. Please circle the overall severity rating for this usability factor:**

| No Usability Problem | Cosmetic Problem Only | Minor Usability Problem | Major Usability Problem | Usability Catastrophe |
|---|---|---|---|---|
| 0 | 1 | 2 | 3 | 4 |

**III. If you have other comments, please specify:**
