## Supplementary material for "Designing a Substance Misuse Data Dashboard for Overdose Fatality Review Teams": ONLINE DATA SUPPLEMENT - Appendix C: Phase 1 Interview Results

| **Theme** | **Subtheme** | **Quote** | **Frequency** |
| --- | --- | --- | --- |
| Challenges | Reliance on manual processes | "Trying to chase after the police officer to get me the reports that I need, or the district attorney to get back to me on any court cases that they might have access to. That's kind of where the burden is, because more often than not I have to send kind reminders that, hey, I'm still looking for this information. And I understand they're busy, too, but you know that information does help us understand things better." (Record 7) | 90% (9) |
|  | Data format and technological issues | Siloed sources  "The biggest thing that's maybe the most cumbersome, we'll use that word again, is just the data collection process. It's very, very involved. There's a lot of different places that data can be gathered, but figuring out where all those places are, how to access all those data points, right? It's very cumbersome.” (Record 10) | 40% (4) |
|  |  | Data format issues  “The new changes, so they recently updated at the national level database and it's made it a lot more time consuming and kind of difficult for our partners to fill out… the questions and answers, especially for the medical examiner, have gotten really lengthy, so it is kind of frustrating trying to find the right boxes to mark.” (Record 2) | 70% (7) |
|  |  | Technological issues  “I'm glad that we have REDCap, but that system can be hard to navigate sometimes. And it's really hard for more than one person to be working in there at a time, so pretty much everything falls under one person when it comes to like running the meeting and then putting in the recommendations and the case data” (Record 1) | 100% (10) |
|  | Mental burden | “Sometimes looking into individuals' lives more than others, or finding more personal information about the individuals whose cases we're reviewing can lead to increased mental workloads for me, like, you know, learning that they had children, or certain things. That certainly adds to the you know humanity of the case review, but also to the realization that you know this, you know, person left people behind or things like that. So, the more that I find, I guess, on my own search can increase the mental workload for me personally.” (Record 3) | 90% (9) |
|  | Time Pressure | “Lots of times for overdose fatality review, and this is true for me also, this is one part of my job, right? It's part of my FTE, it is not my full FTE, so there are directions that I'm pulled for other projects that can sometimes limit the time that I have available to work on this…It's hard to fit this kind of work in for half hours here and 45 minutes there.” (Record 8) | 90% (9) |
|  |  | "OFR can take a lot more time than people realize, and that can be really hard and a reason for why people don't form an OFR, or why they're only meeting quarterly cause we just don't have the capacity to be able to do as much as we would like, just with the time constraints." (Record 1) |  |
| Desired Future State | Easier access to current data sources | "I think something that local public health and us specifically have always sort of struggled with is being able to keep up with that data… having it be a little easier for us to get local data and more quickly, that is an issue that is huge for us. We see things, and we hear about these trends, but we don't necessarily always know that that's happening until all of a sudden, it's like, hey, we're seeing this, you know, all across the county, and is that something we could have caught sooner had we been able to access that data quicker." (Record 1) | 50% (5) |
|  | More data sources | Healthcare data  "Well, always OFR teams, us included, would like more access to data. So I think one of the challenges that all of our teams face is that we don't have any treatment data… it is very difficult to review a death from overdose, a case review and not know whether the person accessed treatment or not." (Record 8) | 70% (70) |
|  |  | Criminal justice data  "It can be a little bit frustrating that some of our information is missing just because busy partners don't have time to look in their systems. So, it's primarily law enforcement, criminal justice contacts that are often missing." (Record 2) | 50% (5) |
|  | Increased collaboration and support | “It's so silly that all the counties in Wisconsin have to do this all ourselves, and I realize that that's just how Wisconsin is organized, and we all have different coroners and medical examiners. But yeah, it's just the fact that everyone's having to basically do the same thing.” (Record 13) | 80% (8) |
|  |  | “I think that there is definitely space in our team for collaboration. I think because this has kind of been started by health departments, a lot of teams kind of see it as something that the Health Department does… And so, I think that there's a lot of room for expansion there that, I think, would also help us kind of stabilize the workflow and identify some of those needs for long-term sustainability.” (Record 1) |  |
