## Supplementary material for "Designing a Substance Misuse Data Dashboard for Overdose Fatality Review Teams": ONLINE DATA SUPPLEMENT - Appendix D: Phase 2 Interview Results

| **Theme** | **Subtheme** | **Quote** | **Frequency** |
| --- | --- | --- | --- |
| Benefits | Improving data access | Expanding access  “A lot of the data that's in here is something that's not necessarily accessible to every county.  I think it would provide an important data source for those counties that maybe don't have their own dashboards or don't have epidemiologists on their staff to help do this type of analysis.” (Record 1) | 100% (7) |
|  |  | Central data access  “That's helpful in just kind of like streamlining it all in one spot, because the data that I do get like from emergency department visits, I have to go through every single entry, and then like, figure it out from there. So, this is quite nice to just have it in one spot where I can look at it, write it down, we're done.” (Record 3) | 71% (5) |
|  | Streamlining workflow | “I really think the timeline is pretty cool for like, for during the review… timelines were something that we incorporated into every case, and it is a lot of time to prepare those if you're doing it manually. So, I thought that was really cool, to see the ability to hover over and get more detail and things like that.” (Record 5) | 86% (6) |
|  | Dashboard structure and organization | Easy to navigate  “I feel like a great emphasis has been placed on trying to make it as user-friendly as possible, and so, there's not a lot of opportunity for it to become frustrating.” (Record 12) | 100% (7) |
|  |  | Filters  “I think that the dashboard was honestly pretty amazing in terms of the filters that you can put in there. Other dashboards I've seen in the past, you're pretty limited in how you can filter the information. You know, you might be able to filter by like race, or age, or gender, but to be able to also have, like what drugs were present, again, fatal versus nonfatal, just those layers, I really appreciated.” (Record 11) | 71% (5) |
|  | Supporting outreach | “My team also works on direct service in harm reduction syringe services program, working directly with people who use drugs, and outreach Narcan® training, those things. I really love that kind of predictive analytics modeling especially at the zip code level, because I think that would be extremely helpful for us or some of the partners that we work with who are in more of an outreach space” (Record 11) | 71% (5) |
| Areas for improvement | Add desired data | “So having social determinants of health will be really helpful, I think. Especially when doing an OFR, because that's the stuff that's hard to get right? And then like I said, behavioral health information” (Record 3) | 86% (6) |
|  | Accurate, representative, and current data | “I like the way that it's laid out, but because it's only people who are going to the ED, I don't know that we can draw conclusions about folks that are at risk of overdose, generally speaking…So, it would depend on how the data looks once populated with actual data for the county, to help us see how well it's aligning with what we know for overdoses when we're looking at all EMS data (which we have access to in our county's dashboard), and then with the fatalities (which we also have access to in the county dashboard).” (Record 1) | 71% (5) |
|  |  | Time lag  “If it takes an extremely long amount of time to get the data, and it's not updated regularly…But I'm used to a huge lag because I work with these other systems where it's gonna take you a year, sometimes 2 to really get information you need. So, if there's any improvement over that, I couldn't see why I wouldn't use it quite honestly.” (Record 11) |  |
|  | Technological suggestions | Exporting data  “I'm thinking more so in terms of practical applications for OFRs. I saw that it generated the timelines and all of that data, and I'm wondering if that's something that could be exported or downloaded, or if we would have to, you know, have 2 different screens, and just manually copy, because that would be a barrier.” (Record 4) | 43% (3) |
|  |  | Other technological suggestions  “So, there's very limited availability to do crosstabs. I realize if you click on, you know, a population of interest, then the other graphs do adjust so you can see maybe just 2 by 2 crosstabs. But, and I realize for confidentiality purposes, you know, people can't have unlimited availability to like filter and crosstab, but I do think there's some limitations in the fact that folks can't do some basic crosstabs by age, sex, and race/ethnicity.” (Record 10) | 57% (4) |
